# Exhausted T cell phenotypes in disseminated coccidioidomycosis

**DOI:** 10.64898/2026.01.31.26345287

**Authors:** Gregory D. Whitehill, Alexis V. Stephens, Timothy J. Thauland, Miguel A. Moreno Lastre, Matthew M. Tate, Sinem Beyhan, Royce H. Johnson, George R. Thompson, Maria I. Garcia-Lloret, Manish J. Butte

**Author notes:** Corresponding author: Manish J. Butte, 10833 Le Conte Ave, MDCC Building, Room 12-430, Los Angeles, CA 90095.

## Abstract

Coccidioidomycosis presents clinically as a spectrum ranging from self-limiting Uncomplicated Valley Fever (UVF) in most cases to life-threatening Disseminated Coccidioidomycosis (DCM) in rare individuals. A few patterns of immunologic deficits allowing for dissemination have been identified, though the specific defects in most individuals with DCM remain undefined. We hypothesized that chronic antigen exposure in DCM engenders a state of T cell exhaustion.

From a cohort of over 300 subjects with confirmed diagnoses of coccidioidomycosis, circulating T cell phenotypes were characterized via flow cytometry and *Coccidioides*-specific T cell responses were measured using the Activation-Induced Marker (AIM) assay.

Male sex was significantly associated with disseminated disease (odds ratio 2.5; 95% CI: 1.5 – 4.0). 52% of subjects showed Coccidioides-specific T cell responses in our AIM assay. We noted a significant difference in subjects sampled in the first year of diagnosis, where only 8% of DCM subjects had T cell responses during this time, as compared to 44% of UVF subjects (p = 0.04). Among DCM patients with detectable AIM responses, CD4+ T cells demonstrated an exhausted phenotype with elevated PD-1 expression compared to UVF subjects. In vitro PD-1 blockade augmented IFNγ production in most tested DCM subjects.

These findings suggest that dissemination may occur in some individuals during a period of impaired antigen-specific T-cell activity. Importantly, these responses can be augmented in vitro by PD-1 blocking antibodies, supporting further study of immune checkpoint therapy as an adjunct to antifungal treatment in disseminated coccidioidomycosis.

## Introduction

Coccidioidomycosis comprises clinical presentations ranging from mild to catastrophic in patients who appear to be otherwise healthy, suggesting that yet unappreciated immunologic deficits may contribute to severe cases.^1^ Coccidioides is acquired by inhalation of environmental conidia, which then propagate in the host as yeast-like spherules. More than half of individuals exposed to Coccidioides develop lifelong protective immunity without recognizable illness.^2^ Symptomatic disease most often presents as uncomplicated Valley Fever (UVF): a self-limited pneumonia. Complicated pulmonary coccidioidomycosis (CPC), characterized by cavitation, respiratory failure, or persistent pulmonary disease despite treatment occurs less frequently and is associated with immunocompromising conditions such as diabetes and advanced age.^3,4^ Extrapulmonary disseminated coccidioidomycosis (DCM) is the rarest and most severe presentation, occurring in only ∼1% of cases and often involving lymphatic tissues, the musculoskeletal system, skin, or the meninges.^5^ Non-meningeal DCM is often chronic, requiring three or more years of antifungal therapy to achieve sustained remission, while meningitis is almost universally fatal without lifelong treatment.^6,7^ Even with antifungal therapy, mortality for severe pulmonary or disseminated Coccidioides requiring ICU admission approaches 50% in some contemporary cohorts.^8^ The burden of Coccidioidomycosis is increasing with recent yearly incident rates estimated at 206,000 – 360,000.^9^ Immunomodulatory therapy is a promising approach to rescue antifungal refractory disease and hasten recovery in severe cases.

Type 1 T-helper (Th1) responses are central for control of Coccidioides infections, as seen through mouse models and vaccine studies.^10^ Th1 cells direct delayed-type hypersensitivity (DTH) reactions and augment the phagocytic killing of Coccidioides through the action of their key cytokine interferon gamma (IFNγ). Patients with coccidioidomycosis demonstrating erythema nodosum, a DTH-mediated panniculitis, have favorable outcomes (e.g., UVF) even among high-risk patient populations^11^. On the other hand, impaired Th1 function leads to worse outcomes. Inborn errors of immunity affecting pathways critical for Th1 induction and effector function (such as IL-12 or IFNγ signaling) are overrepresented in patients with DCM.^12–14^ Lymphocyte-suppressive medications for solid organ transplant or rheumatologic disease are also associated with increased risk for DCM.^12,15^ Patients with DCM are less likely to present with pulmonary nodules, suggesting an impaired granulomatous response at the time of infection in these more severe presentations.^6^

Reliable detection of Coccidioides-specific T cells has proven challenging but prior studies suggest that the magnitude of Th1 responses is lower in severe disease presentations. In the early to mid-20^th^ century, skin testing to measure DTH reactions against Coccidioides conidia or spherule-derived antigens was purportedly prognostic, with robust skin test responses predicting convalescence and anergic responses predicting disseminated disease and relapse.^2,16^ More recent studies however have found no clear association between skin test results and disease severity or outcome, at least possibly due to heterogeneities in the skin test reagents.^6,17^ In vitro studies, primarily employing the *C. posadasii* spherule-derived T27K antigen, have shown that patients with UVF exhibit greater T cell responses than those with DCM as measured by proliferation (^3^H-thymidine uptake), IFNγ production, and CD69 expression.^18–21^ The disparity in T cell responses between disease states may be most prominent during the first six to twelve months of infection, during which time patients with DCM interestingly mount exaggerated inflammatory responses with notable elevation of IL-10 and IL-6.^20,22,23^ DCM is also associated with high-titer Coccidioides-specific antibodies which wane slowly with treatment.^24^ Thus while patients with DCM mount robust immunity, the response appears biased towards exaggerated humoral inflammation but impaired T cell activity.

A state of T cell “exhaustion” arises in chronic disease states where there is prolonged exposure to antigen, such as in chronic infections or cancers. Chronic T cell receptor signaling imposes permanent epigenetic changes that impair cytokine production and cytotoxic killing.^25,26^ While protective against autoimmunity or excessive tissue injury from exaggerated immune responses, exhaustion also inhibits beneficial cellular immunity. Monoclonal antibodies that block the inhibitory receptor PD-1 can transiently restore the effector functions of exhausted T cell responses, and have become the standard of care for several cancers.^27^ Development of granulomata in fungal infections arise from local Type 2 immune responses^28^ and allow fungi to persist in a latent form for years or decades.^29^ The impaired Th1 responses of early DCM may promote T cell exhaustion by establishing a long-lasting antigenic depot within the immunosuppressive granulomatous milieu. Mouse models demonstrate that Coccidioides induces expression of PD-L1 on antigen presenting cells, suggesting that targeting the PD-1/ PD-L1 pathway may have therapeutic merit.^30^ PD-1 blockade has shown promise in mouse models of dimorphic fungal infection and several human cases of invasive mold infection.^31–37^

We hypothesized that the chronic disease state of DCM, characterized by prolonged exposure to cocci antigens (as potentially evidenced by prolonged elevation of antibody titers) and dysfunctional T cell responses may create an environment that fosters T cell exhaustion. In this study, we characterized peripheral T cell phenotypes in a large cohort of subjects with a range of Coccidioides disease and measured Coccidioides-specific T cell responses by Antigen-Induced Marker (AIM) assay. In DCM, T cell responses correlated with later stages of disease and increased expression of exhaustion markers. Our findings suggest that in some patients with DCM, Coccidioides-specific T cells are exhausted and can be augmented by PD-1 blockade.

## Results

### Cohort Characteristics

Three hundred and ten subjects with confirmed diagnosis of coccidioidomycosis were recruited. Clinical disease category and demographics are reported in Table 1. Age, sex, self-identified race and ethnicity (SIRE), and date of initial Coccidioides diagnosis were provided by subjects at recruitment. Coccidioidomycosis severity for each subject was categorized using the five-level numeric classification described by Krogstad and colleagues (1, asymptomatic; 2, uncomplicated pulmonary; 3, complicated pulmonary; 4, non-meningeal disseminated; 5, meningitis).^1^ Complicated pulmonary coccidioidomycosis (CPC), non-meningeal disseminated coccidioidomycosis (NMDCM), and Coccidioides meningitis (CM) were further stratified into subcategories. CPC was heterogenous, comprising approximately equal proportions of simple cavitary disease (3A, 41%) and persistent symptoms despite six months of therapy (3C, 46%) with few cases of respiratory failure (3D, 10%) or fibrocavitary disease (3B, 3%). For NMDCM, most had dissemination to extrapulmonary organs or deep tissue sites (4B, 88%) while only a few showed dissemination to skin only (4A, 12%). Most subjects with CM had isolated meningeal disease (5A, 85%), while fewer had additional dissemination to skin (5B, 4%) or deeper tissue sites (5C, 11%).

**Table 1:** Cohort and Demographics.

| Disease Category | 1 (ASX) | 2 (UPC) | 3 (CPC) | 4 (NMDCM) | 5 (CM) |
| --- | --- | --- | --- | --- | --- |
| Assignment | UVF | UVF | n/a | DCM | DCM |
| N (%) | 17 (6%) | 95 (31%) | 59 (19%) | 67 (22%) | 72 (23%) |
| Age (y) | 38 (29-40) | 43.0 (33–57) | 49.0 (36–57) | 50.0 (36–58) | 52.0 (35–58) |
| Months since Dx | 12 (5-27) | 7 (3-51) | 34 (14-64) | 30 (15-73) | 77 (38-144) |
| Sex |  |  |  |  |  |
| Female | 11 (65%) | 47 (50%) | 24 (41%) | 15 (22%) | 27 (38%) |
| Male | 6 (35%) | 48 (51%) | 35 (59%) | 52 (78%) | 45 (63%) |
| SIRE |  |  |  |  |  |
| Hispanic/Latino | 16 (94%) | 66 (71%) | 33 (56%) | 34 (52%) | 49 (69%) |
| White |  | 10 (20%) | 16 (27%) | 18 (27%) | 16 (23%) |
| Black |  | 2 (2%) | 4 (7%) | 8 (12%) | 4 (6%) |
| Asian | 1 (6%) | 6 (7%) | 4 (7%) | 4 (6%) | 2 (3%) |
| Pacific Islander |  |  | 1 (2%) | 2 (3%) |  |
| American Indian |  |  | 1 (2%) |  |  |
| N/A |  | 11 |  | 1 | 1 |
| Subcategory |  |  | 3A: 24 (41%)<br>3B: 2 (3%)<br>3C: 27 (46%)<br>3D: 6 (10%) | 4A: 8 (12%)<br>4B: 59 (88%) | 5A: 61 (85%)<br>5B: 3 (4%)<br>5C: 8 (11%) |

ASX, asymptomatic; UPC, uncomplicated pulmonary *Coccidioides*; CPC, complicated pulmonary *Coccidioides*; NMDCM, non-meningeal disseminated *Coccidioides*; CM, *Coccidioides* meningitis. UVF, uncomplicated valley fever; DCM, disseminated *Coccidioides*. SIRE; self-identified race and ethnicity. Dx, diagnosis.
For Age and Months since diagnosis, median (IQR) are presented within each category.
For SIRE, % includes only subjects who responded (N/A excluded).
Subcategories: 3A, pulmonary disease with simple cavitation; 3B, fibrocavitary lung disease; 3C, persistent pulmonary disease for >6 months despite therapy; 3D, respiratory failure. 4A, dissemination to skin only; 4B, dissemination to sites other than skin. 5A, meningitis without other extrapulmonary dissemination; 5B, meningitis with dissemination to skin but no other sites; 5C, meningitis with dissemination to skin and/or other organs.

We investigated whether demographic variables were associated with severe disease states by consolidating subjects into two broader groups: uncomplicated valley fever (UVF, categories 1 and 2) and disseminated coccidioidomycosis (DCM, categories 4 and 5). CPC were excluded from these analyses due to the clinical heterogeneity of this group which suffered more complicated syndromes than the self-limited UVF, yet without extrapulmonary dissemination. Male sex was significantly associated with DCM (Fisher exact test, p = 0.007; odds ratio (OR) [95% CI] = 2.5 [1.5 – 4.0], p < 0.001, **Figure S1**) as other studies also observed.^2^ Prior studies suggest that persons identifying as Hispanic, Black, or Asian have increased risk for disseminated coccidioidomycosis relative to those identifying as White.^38,39^ However, our cohort showed that SIRE of Hispanic or Latino was associated with a decreased risk for DCM (OR [95% CI] = 0.4 [0.2 – 0.7]) while risk was increased among non-Hispanic White (OR = 3.1 (1.5 – 6.2) and Black (OR = 4.2 (1.2 – 22)) but not Asian subjects (**Figure S1**). Regarding age, DCM subjects were older than UVF at recruitment (median [IQR] = 50 years [34 – 58] vs 43 [33 – 56], p = 0.05 by Mann-Whitney) but age at diagnosis was not different (41 years [28 – 50] vs 40 years [20 – 53], p = 0.6 by Mann-Whitney). Notably, subjects with UVF were recruited significantly more proximally to their dates of diagnosis than those with DCM (9 months [3 - 46] vs 57 months [20 - 129], p < 0.0001 by Mann-Whitney). Taken together, these data show our cohort comprises a typical and diverse spectrum of demographics and clinical phenotypes.

### T Cell Phenotype in DCM Suggests Exhausted Immune State

We assessed whether biomarkers of peripheral T cell differentiation and activation differed between the coccidioidomycosis disease severities. By flow cytometry we measured naive and memory subsets (Naïve, EM, CM, TEMRA) and activation/ exhaustion markers HLA-DR, PD-1, and CD57 in peripheral blood CD4+ and CD8+ T cells (**Figure 1, S2**).

**Figure 1:**
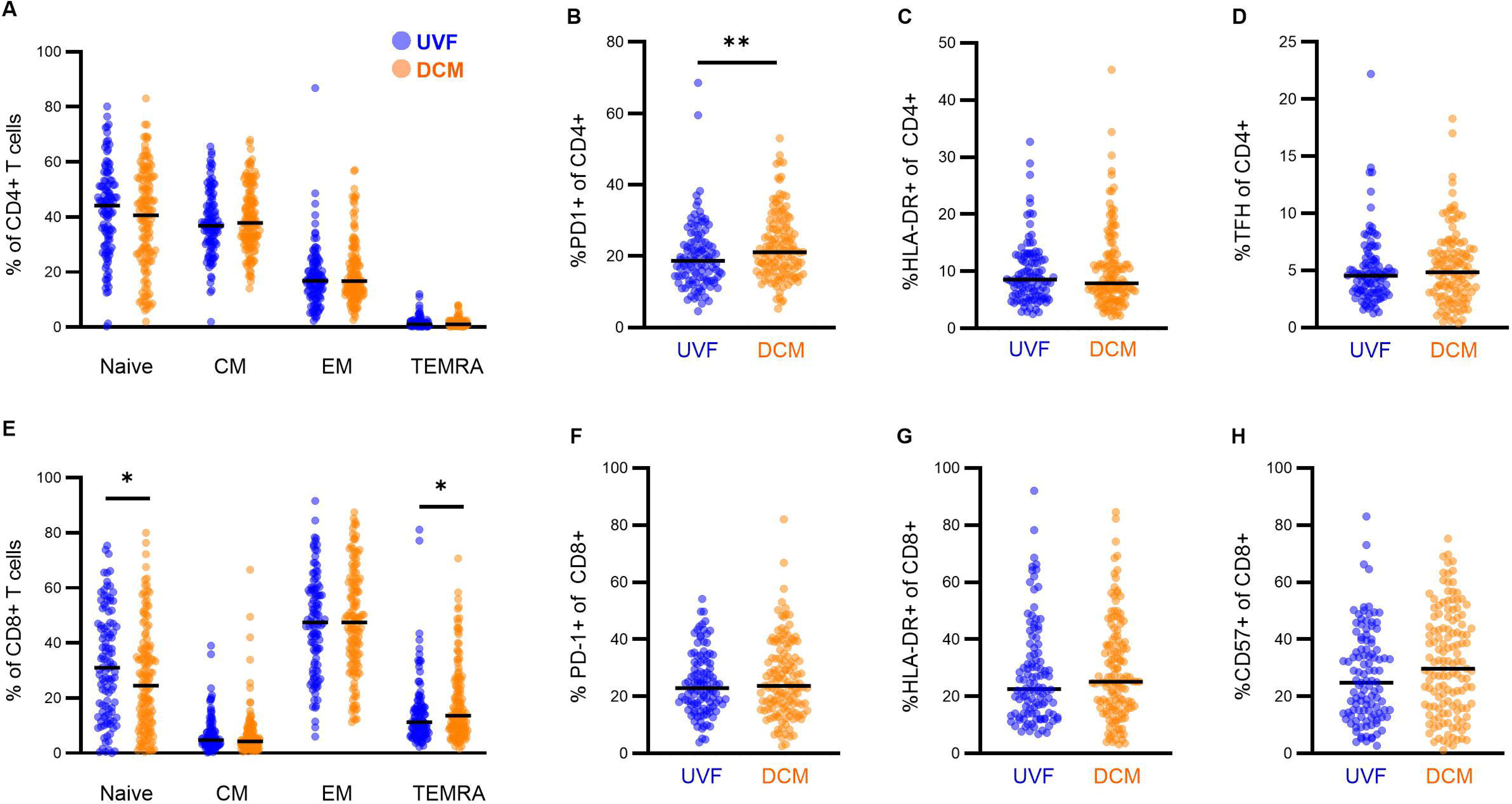
PBMC T cell Phenotype of uncomplicated Valley fever (UVF) and disseminated coccidioidomycosis (DCM). PBMCs from patients with UVF (blue, N = 108) and DCM (orange, N = 130) were phenotyped by flow cytometry. (**A, E**) Naïve (CD45RA+CCR7+), central memory (CM, CD45RA-CCR7+), effector memory (EM, CD45RA-CCR7-), and TEMRA (CD45RA+CCR7-) subsets of (**A**) CD4+ and (**E**) CD8+ T cells. (**B - D**) Percentage of (**B**) PD-1+, (**C**) HLA-DR+, and (**E**) T-follicular helper (TFH, PD1+CXCR5+) CD4 T cells. (**F - H**) percentage of (**F**) PD-1+, (**G**) HLA-DR+, and (**H**) CD57+ CD8 T cells. Statistical comparisons by Mann-Whitney U-test, unadjusted. *, p < 0.05; **, p < 0.01.

CD4 T cell memory subsets were not significantly different between subjects with DCM or UVF, however the DCM group showed a trend towards decreased CD4+ naïve T cells compared to UVF (median [IQR] = 41% [26 - 54] vs 44% [30 - 53], p = 0.2 by Mann Whitney, **Figure 1A**). PD-1 expression on CD4 T cells was significantly greater in the DCM group relative to UVF (21% [16 - 28] vs 19% [13 - 24], p = 0.007, **Figure 1B**). PD-1 is expressed on T cells shortly following TCR stimulation, and can demonstrate an exhausted T cell phenotype at steady state months to years after acute infection.^40,41^ Activated T cells additionally express HLA-DR, which is associated with a replicating effector state.^42,43^ HLA-DR expression was not significantly different between the groups, trending lower in DCM compared to UVF (8% [6 - 13] vs 9% [6 - 12], **Figure 1C**). PD-1 is also expressed on circulating follicular helper (TFH) CD4+ T cells and facilitates productive interactions between T and B cells.^44^ Coccidioides-specific antibody titers ore often dramatically elevated in patients with DCM. To assess whether the increased PD-1 expression in this group was driven by an expanded TFH subset we measured CD4+PD-1+CXCR5+ circulating TFH cells, which were not significantly different between DCM and UVF (5 [3 - 7] vs 5 [3 - 6], **Figure 1D**).

CD8 T cell memory populations trended similarly to CD4 T cells. Naïve CD8 T cells subsets were smaller in DCM compared to UVF (24% [11 - 38] vs 31% [15 - 46], p = 0.03) and terminally differentiated TEMRA populations were larger (9% [2-14] vs 7% [2-11], p = 0.04) (**Figure 1E**). CD57 expression (30% [16 - 46] vs 25% [14 - 39], p = 0.05, **Figure 1H**) and to a lesser extent PD-1 expression (24% [16 - 34] vs 23% [17 - 32], p = 0.7, **Figure 1G**) on CD8 T cells also trended higher in DCM than UVF.

T cell phenotypes suggested a more differentiated and exhausted state in DCM relative to UVF with significantly increased PD-1 expression among CD4 T cells, decreased naïve T cells, and a trend towards increased PD-1+ and CD57+ CD8 T cells. Since the two groups also varied by age, sex, and time since diagnosis, we assessed whether these variables confounded our findings. CD4 PD-1 expression showed a positive but statistically insignificant correlation with age in both UVF and DCM, with a stronger correlation among DCM relative to UVF (Spearman r = 0.098 vs 0.026) (**Figure S3A, B**). We did not identify significant correlations between CD4 PD-1 expression with time since diagnosis or sex either (**Figure S3C - F**). As some of the variance in PD-1 expression could be attributed to subject age, we assessed the variance of all measured phenotypic markers with age by linear regression. Across the entire cohort, age associated similarly with CD4 PD-1+ cells (r^2^ = 0.014) compared to CD4 HLA-DR+ cells (r^2^ = 0.023), while other subsets such as CD4 CM (r^2^ = 0.11), CD8 Naïve (r^2^ = 0.24), CD8 HLADR+ (r^2^ = 0.074), CD8 CD57+ (r^2^ = 0.061) showed a much stronger association with age (**Figure S4**). These results suggest that age does not sufficiently explain the variance in CD4 T cell PD-1 expression between DCM and UVF. We hypothesized that this phenotypic difference may reflect an exhausted immune state, either due to persistent T cell stimulation from the chronic nature of disseminated coccidioidomycosis or other host factors which we did not assess. In cancer, certain markers in pre-treatment PBMC have been associated with favorable responses to checkpoint inhibitor therapy, including CD57 expression on CD8 T cells and increased CD4 memory populations relative to naïve.^45,46^ We next sought to compare Coccidioides-specific T cell responses between UVF and DCM.

### AIM assay for measurement of Coccidioides-specific T cell responses

Coccidioides-specific T cell responses were measured by AIM assay.^47^ PBMCs were stimulated separately with formalin-killed spherules (FKS) or formalin-killed conidia (FKC) derived from the *C. immitis* RS strain. Activated CD4+ T cells were identified by co-expression of both CD69 and CD134 (OX40), and activated CD8+ T cells by both CD69 and CD137 (4-1BB) (**Figure S5**). AIM assay allows for the identification of diverse populations of antigen-specific CD4 and CD8 T cells specific for linear peptide antigens (e.g. SARS-CoV-2 and HCV) and more complex antigen preparations such as lysates of mycobacterium tuberculosis and Trypanosoma cruzi.^48–51^ T cells are reported to express AIM markers both in response to TCR engagement with cognate peptide-MHC complexes and, when cultured over longer periods, antigen-independent ‘bystander’ activation by stimulatory cytokines.^52^ We tested whether AIM responses to FKC and FKS were TCR dependent by stimulating PBMCs in the presence of MHC-I and MHC-II blocking antibodies (**Figure 2A, B**). Both CD4 and CD8 T cell responses were fully suppressed by MHC-II blockade but not MHC-I blockade, suggesting that FKS- and FKC-responsive CD4+ T cells were antigen-specific while bystander activation drove CD8+ T cell responses. This finding is consistent with extracellular antigen processing predominantly utilizing endosomal pathways and presentation on MHC-II molecules. Because of this finding and because the role of helper T cells is much better defined in fungal immunity, we therefore directed our focus to CD4+ T cells.

**Figure 2:**
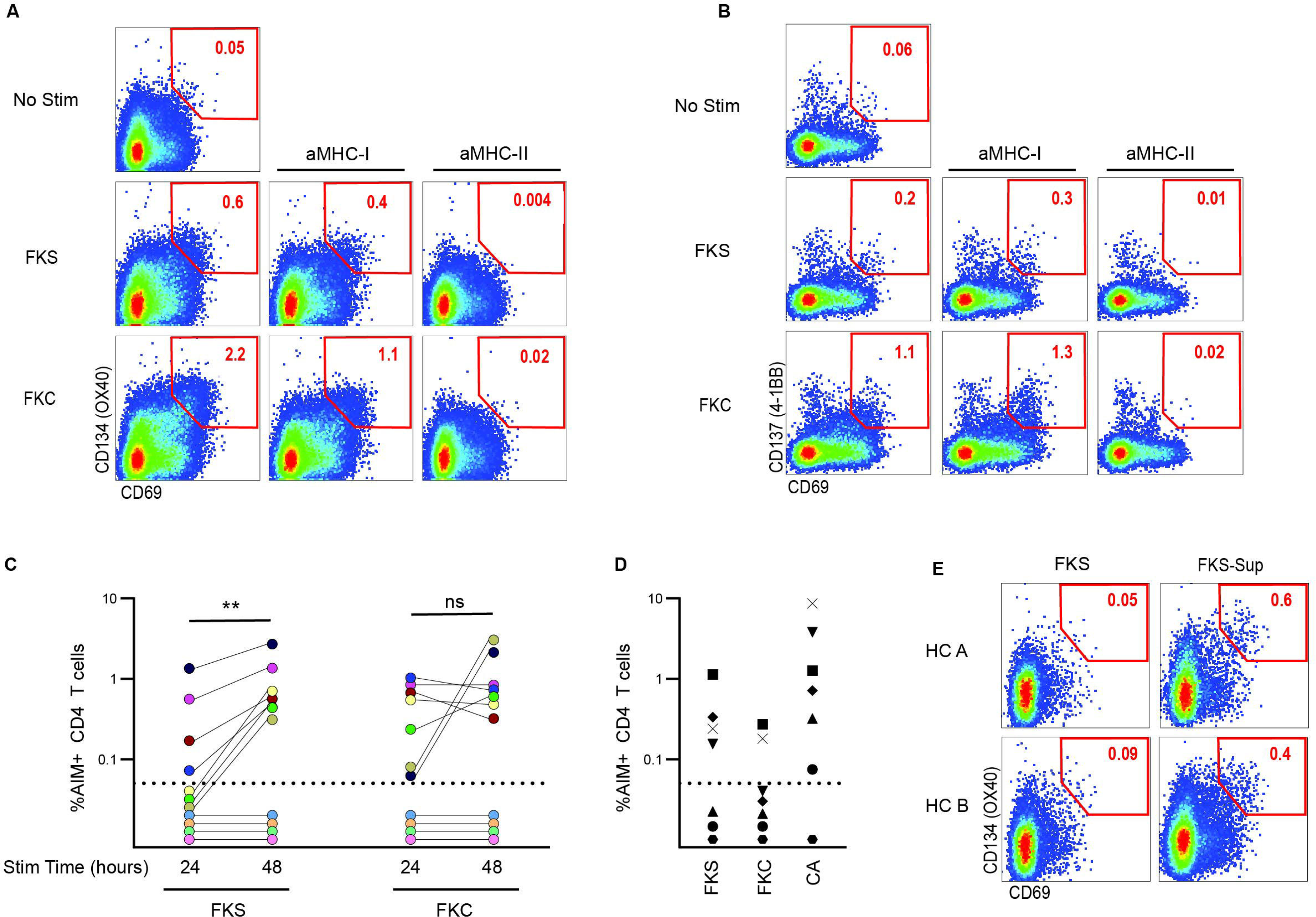
AIM assay to identify Coccidioides-specific T cell responses. (**A**, **B**) PBMCs were incubated with Coccidioides formalin-killed spherules (FKS), formalin-killed Conidia (FKC), or media alone (No stim) with or without MHC-I blocking antibody (αMHC-I) or MHC-II blocking antibody (αMHC-II) for 24 hours. AIM-positive populations are gated for (**A**) CD4+ T cells (CD69+OX40+) and (**B**) CD8+ T cells (CD69+CD137+). (**c**) PBMCs from 11 subjects with Coccidioidomycosis (mixed UVF and DCM) were simulated with FKS and FKC for 24 and 48 hours. Percent AIM-positive CD4 T cells with unstimulated background subtracted are presented. Colors denote unique subjects. Comparisons by Wilcoxon matched pairs ranked sum test **, p < 0.01. (**d**) PBMCs from 7 healthy control donors (HC) stimulated with FKC, FKS, or candida antigen (CA) for 24 hours. Symbols denote unique subjects. For (**C**) and (**D**), values below dotted line (0.05%) are negative. (**E**) PBMC from two healthy controls (HC A and HC B) were stimulated for 48 hours with FKS alone or supernatant collected from cultures of Coccidioides subject PBMCs stimulated with FKS for 48 hours (FKS-sup). AIM-positive population of CD4+ T cells is gated.

To assess the optimal stimulation period for these antigens, we compared T cell responses against FKS and FKC over 24-hour and 48-hour incubation periods (**Figure 2C**). FKS-specific responses were greater at 48 hours stimulation than 24 hours (p = 0.008, Wilcoxon matched pairs ranked sum test), while FKC responses were not significantly different between the two timepoints. Seven of the eleven subjects (64%) demonstrated a detectable response to any antigen across both timepoints, and all of these showed positive FKC-specific responses at 24 hours. These results show that 24-hour stimulation with FKC was adequate to detect cocci-specific T cell responses in over half the subjects, and FKS required a longer stimulation period.

To assess the specificity of FKS and FKC-specific responses, we examined the AIM assay in PBMCs from endemic-region healthy control donors (HC, N = 7) with an unknown history of Coccidioides exposure (**Figure 2D**). Four HC (57%) showed a positive response to FKS, while two (29%) responded to FKC. Interestingly, the four HC with FKS-specific responses also showed high T cell responses to stimulation with candida antigen (CA). In vitro T cell cross-reactivity between Coccidioides with other endemic mycoses including Histoplasma and Blastomyces has been described, but whether Coccidioides-specific T cells cross-react with Candida is not known.^53^ We then attempted to determine whether the increased FKS-specific responses at 48 hours were driven by delayed T cell activation or bystander activation. PBMCs from two HC were stimulated with FKS alone for 48 hours or with supernatant (FKS Sup) collected from AIM-positive subjects stimulated with FKS for 48 hours. FKS Sup-stimulated HC PBMCs showed a greater percentage of AIM+ CD4 T cells than FKS alone (**Figure 2E**). This result showed that soluble factors produced by FKS-stimulated cells could augment T cell responses over extended incubation periods. Therefore, while the extended 48-hour stimulation period was necessary to reliably detect FKS-specific responses, some of these responses could also accumulate the effects of bystander T cell activation.

The finding of bystander effect and possible cross-reactivity with other fungal antigens does not diminish the relevance of spherule-specific T cell responses. Coccidioides is transmitted as conidia form but propagates within infected hosts as spherules, and these two forms demonstrate unique expression patterns of their transcriptomes and proteomes.^54^ The degree of antigenic overlap between Coccidioides spherules and conidia relative to MHC-presented peptides and the T cell repertoire which recognizes them has not been characterized. We therefore concluded that the presence or absence of Coccidioides-specific T cell responses could be adequately assessed by either FKC at 24 or 48 hours of stimulation, or FKS at 48 hours stimulation, though the absolute magnitude of response may differ by the method employed.

### T cell responses are impaired early in Disseminated Coccidioides

We surveyed Coccidioides-specific T cell responses by AIM assay in 161 subjects from various disease categories across our cohort (**Figure 3A**). Positive responses were detected in 52% of subjects (N = 84 of 161). By disease category, the response rates were 50% for asymptomatic disease (ASX, 5 of 10), 51% for uncomplicated pulmonary (UPC, 26 of 51), 59% for complicated pulmonary (CPC, 13 of 22), 40% for non-meningeal disseminated cocci (NMDCM, 19 of 47), and 68% for cocci meningitis (CM, 21 of 31). Given the low response rate, we sought to validate that the AIM assay correlated with functional T cell responses. We tested 22 AIM-positive subjects and 14 AIM-negative subjects for production of IFNγ, IL-4, and IL-17A in response to stimulation with cocci antigens (**Figure 3B, S6**). FKS and FKC provoked similar responses, and cytokine production was almost universally absent in subjects who lacked an AIM response.

**Figure 3:**
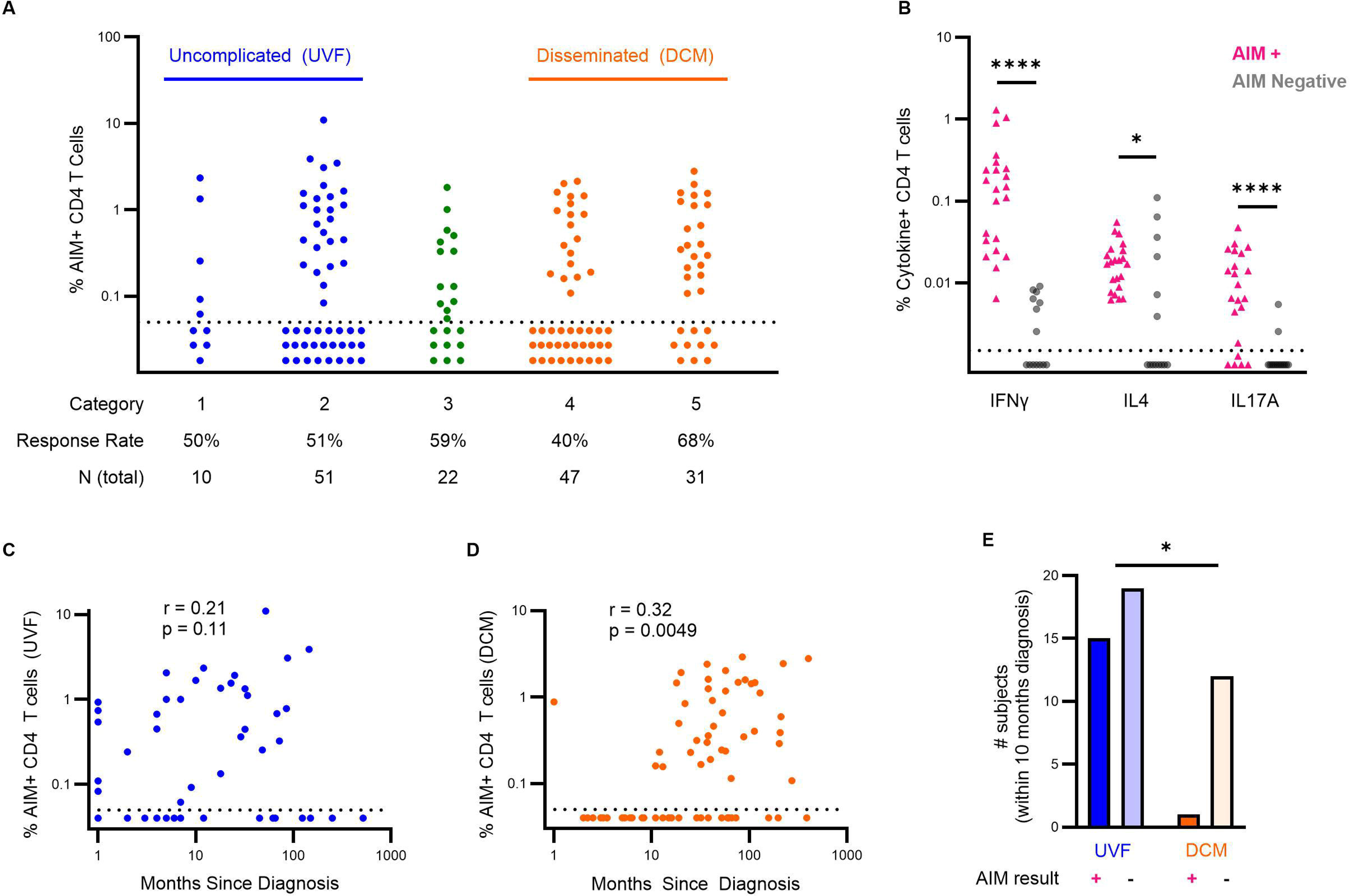
Coccidioides specific T cell responses for 161 subjects by disease category. AIM responses stratified by Coccidioides disease category. Limit of detection = 0.05%, values below dotted line are negative. (**B**) IFNγ, IL-4, and IL-17 production of CD4 T cells stimulated with FKS or FKC from subjects with positive Coccidioides-specific T cell responses by AIM assay (AIM+, pink, N = 22) or no response (AIM negative, grey, N = 14). Comparisons by Wilcoxon signed rank test. (**C**, **D**), Spearman correlation of Coccidioides-specific T cell responses by AIM and time since diagnosis for (**C**) UVF and (**D**) DCM. (**E**) Number of subjects with UVF or DCM within ten months of diagnosis with positive or negative Coccidioides-specific T cell responses by AIM assay, Fisher exact test. *, p < 0.05; ***, p < 0.005.

Most of our subjects with DCM were recruited years after their initial diagnosis and those with UVF were mostly recruited within one year. To investigate whether the proximity of blood sampling to diagnosis affected T cell responses, we compared AIM responses by time since Coccidioides diagnosis. The result revealed a strikingly low response rate amongst DCM subjects recruited early in disease (**Figure 3C, D**): only 1 of 13 (8%) DCM subjects within 10 months of diagnosis demonstrated a positive AIM response. In contrast, positive responses were detected for 15 of 34 (44%) UVF subjects within 10 months of diagnosis (p = 0.04, Fisher exact test, **Figure 3E**). These results reveal an almost one-year refractory period of Coccidioides antigen recognition by CD4 T cells for subjects with DCM but not UVF.

We compared AIM responses by age and sex as these variables also differed between UVF and DCM (**Figure S7**). AIM responses did not vary significantly with age for UVF or DCM (**Figure S7A, B**). For both UVF and DCM, a larger percentage of females were AIM responders than males (**Figure S7C, D**). This sex-dependent difference in response rate was more was pronounced in DCM (Female, 68%; Male, 45%) than UVF (Female, 55%; Male, 47%) but was not statistically significant (p= 0.09, 0.6, respectively, by Fisher Exact test).

### Disseminated Coccidioides patients with detectable T cell responses show an exhausted phenotype and respond to checkpoint blockade in vitro

Having identified an apparent deficit in the induction of T cell responses in DCM, we then assessed whether DCM subjects with T cell responses (i.e. ‘AIM-positive’) demonstrated functional T cell exhaustion. Revisiting the PBMC T cell phenotypes presented in Figure 1 among just the AIM-positive subjects, more CD4 T cells expressed PD-1 among AIM-positive DCM subjects compared to AIM-positive UVF subjects (median = 24% vs 20%, p = 0.01 by Mann-Whitney, **Figure 4A**). Interestingly, CD4 PD-1 expression was significantly lower among the AIM-negative DCM group, suggesting that the increased PD-1 expression in DCM may be driven by Coccidioides-specific immune responses (**Figure S8**). We tested whether Coccidioides-specific T cell effector responses could be augmented by checkpoint blockade. PBMC from 13 AIM-positive DCM subjects were stimulated with FKS or FKC in the presence of PD-1 inhibitor or isotype control (**Figure 4B, C**). Ten subjects (77%) showed an increase in CD4 T cell IFNγ production with PD-1 blockade, and in six subjects (46%) the increase was two-fold or greater (**Figure 4C**). Increases in IL-17A production, and to a lesser extent IL-4 production, were observed in some subjects as well. Checkpoint blockade did not boost cytokine production in AIM-negative subjects.

**Figure 4:**
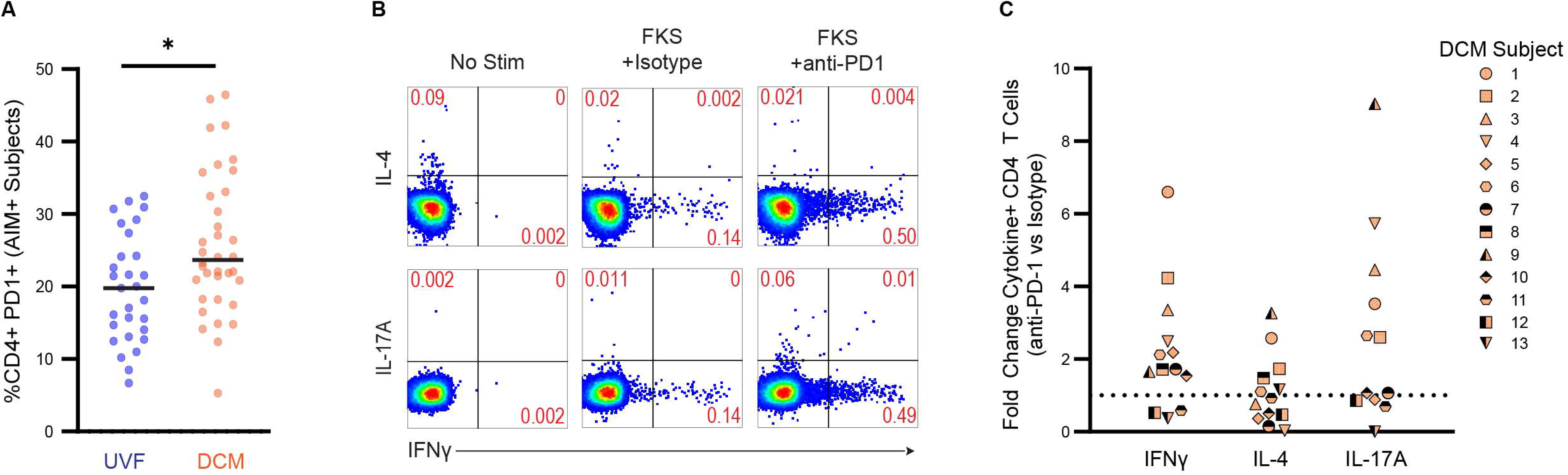
Checkpoint blockade augments Coccidioides specific T cell responses in DCM. (**A**) %PD1+ CD4 T cells among AIM-positive UVF (blue, N = 29) and AIM-positive DCM (orange, N = 36) subjects, comparison by Mann-Whitney U Test, *, p < 0.05. (**B - C**) PBMC from N = 13 AIM-positive DCM subjects were cultured with media alone (no stim), FKS with isotype control antibody, or FKS with anti-PD-1 antibody. IFNg, IL-4, and IL-17A production were measured by intracellular staining and flow cytometry. (**B**) Representative subject data. (C) Fold change cytokine production, each symbol represents one subject.

We attempted to identify phenotypic or demographic correlates which were associated with response to checkpoint blockade. Fold change in interferon gamma production against Coccidioides antigen with PD-1 blockade was compared to phenotypic markers, AIM response, age, and time since infection by linear regression (**Figure S9**). The two highest responders (IFNg fold change 4.2 and 6.6) were both female with CD4 PD-1 expression >40% and the three nonresponders were all male. However, no phenotypic or demographic variable assessed showed significant variation with response to PD-1 blockade.

## Discussion

We investigated peripheral T cell phenotypes and Coccidioides-specific CD4 T cell responses in a large cohort with various manifestations of Coccidioidomycosis. We found that peripheral T cell repertoires in disseminated disease trended towards an exhausted, antigen-experienced phenotype by expanded memory populations, and increased expression of PD-1 on CD4 T cells which was not attributable to age. T cell responses against a novel set of Coccidioides antigens measured by AIM assay showed a nearly one-year refractory period in DCM. The DCM subjects driving phenotypic differences in PD-1 expression relative to UVF were those with detectable Coccidioides-specific T cells, suggesting an association between exhausted phenotypes and Coccidioides-specific immune responses. A subset of subjects with DCM showed boosting of functional T cell responses to Coccidioides antigens by in vitro PD-1 blockade. Our findings support that T cell exhaustion may contribute to T cell dysfunction in some cases of disseminated Coccidioidomycosis and warrants further exploration of adjunct checkpoint inhibitor therapy for severe cases.

In our cohort, male sex was associated with increased risk for disseminated disease, as was non-Hispanic White or Black SIRE. Male sex has been observed as a risk factor for Coccidioidomycosis as well as a broad host of viral, bacterial, and fungal infections. While the increased susceptibility of men to Coccidioidomycosis is not fully understood, testosterone has been described to suppress Th1 differentiation and function, which may partially explain sex differences in disease risk.^55^ African and Filipino ancestry have been consistently associated with more severe Coccidioides disease states in prior studies.^56^ Our cohort recapitulates the increased risk of DCM among individuals identifying as black SIRE, despite representing a minority of subjects (N = 18). We did not specifically elicit Filipino ancestry from subjects in this study. The findings of decreased risk for DCM among Hispanic or Latino subjects and increased risk among non-Hispanic White subjects is contrary to prior findings but may reflect that our cohort is mostly Hispanic or Latino (N = 198 of 310, 64%).

We expected that all immunocompetent patients with coccidioidomycosis would mount a detectable T cell response to at least some *Coccidioides* antigens. However, the history of assessing functional T cell responses in coccidioidomycosis is fraught with two major concerns: inadequate sources of *Coccidioides* antigens and inadequate readouts. For this reason, we introduced the AIM assay here to detect Coccidioides-specific T cell responses with novel antigenic preparations derived from *Coccidioides immitis* RS strain spherules and conidia. Other sources of cocci antigen have been employed over the years with modest-to-poor elicitation of Coccidioides-specific T cell responses. T cell responses to the *Coccidioides posadasii* spherule lysate T27K were assessed by expression of the sole activation marker CD69 and by cytokine production in endemic healthy controls, nonendemic healthy controls, and Coccidioides patients with variable severity of disease.^18,20,57,58^ Though response rates in these studies are not reported, in vitro stimulation with T27K showed 83-85% agreement with Coccidioidin skin testing.^59^ More remote studies in the 1970s also showed that in vitro lymphocyte stimulation with mycelial or spherule derived lysates largely agree with skin test results.^21^

Skin testing (namely, delayed-type hypersensitivity (DTH)) with spherule-derived antigen was originally described to demonstrate >98% sensitivity and specificity for patients with pulmonary coccidioidomycosis, suggesting that in vitro responses to Coccidioides antigens should be much higher than the 54% than we observed in our cohort.^60^ However, more recent studies suggest that in practice, only 55-72% of patients with a Coccidioides diagnosis have a positive DTH.^61,62^ Antifungal therapy reduces fungal burden and may be associated with decreased skin test sensitivities, suggesting that prolonged untreated infection may be necessary to developed detectable T cell responses in some cases.^61^ During acute infection, T cells may also be sequestered to lymph nodes or infected tissue sites and thus not reliably detected in peripheral blood alone. For example, in CM, *Coccidioides*-specific T cells can durably be detected in CSF while absent from peripheral blood.^63^ Peptide antigens are also under development for diagnostic testing. Kala and colleagues recently designed a mix of 108 peptides derived from *Coccidioides* proteins that induce T cell responses in mice and are upregulated during the parasitic growth phase.^64^ However, even this more modern approach did not detect a response in all subjects with coccidioidomycosis. In our hands, the AIM assay in CD4+ T cells passed many important measures of quality, including dependency on MHC-II interactions and correlation with cytokine production. Whether the difficulty in reliably detecting these responses in peripheral blood is due to tissue compartmentalization, anergy, lack of optimized antigen, or inadequate assays warrants further study.

Coccidioides-specific T cell responses within our cohort identified a profoundly low response rate among DCM subjects sampled within 10 months of diagnosis. Similar findings have been reported by Ampel and colleagues, who described decreased T27K-specific T cell responses in DCM compared to UVF during the first 5 and 12 months of infection by CD69 expression and whole blood IFNγ production, respectively.^20,22^ Dissemination most often occurs within 6 months of diagnosis, suggesting that an adequate T cell response during this initial phase of disease is critical to prevent dissemination.^6^ Unfortunately, the low response rate to Coccidioides antigens detected by our assay, while not unexpected in the context of prior studies, is a major caveat which limits decisive interpretation of these results.

Our observation of exhausted T cell signatures in some subjects with DCM invokes CD4 T cell exhaustion, which is less well described than CD8 T cell exhaustion. Initial descriptions of the molecular signatures underlying T cell exhaustion were borne from the study of CD8 T cells in CD4-depleted mouse models of chronic viral infection.^65^ Studies of CD4 T cell exhaustion in this same model showed some overlapping signatures with CD8 T cell exhaustion, but also more heterogeneity in CD4 T cell populations and distinct expression patterns. Relative to CD8 T cells, exhausted CD4 T cells show more sustained high levels of PD-1 expression during chronic infection, and in our cohort PD-1 expression on peripheral CD4 T cells associated with disseminated disease and response to checkpoint blockade.^66^ In cancer models, CD8 T cell exhaustion has also been the primary focus of study due to the lack of MHC-II expression on most tumors and the aim of augmenting cytotoxic responses in this setting. However, CD4 T cells may be important in the response to checkpoint inhibitor therapy as well.^67^ Exhausted tumor-specific CD4 T cells can be found in solid malignancies and show increased functional responses with checkpoint inhibitor therapy.^68^ The role of CD4 T cell exhaustion is more clearly defined in MHC-II expressing tumors such as Hodgkins lymphoma, wherein tumor infiltrating lymphocyte populations are rich in CD4 T cells expressing classical markers of exhaustion (i.e. PD-1 and TOX) and show enhanced antitumor activity with PD-1 blockade.^69,70^ Interpretation of PD-1 expression on CD4 T cells is also muddied by the role of PD-1 in regulating TFH interactions with B cells and the activity of CD4+ regulatory T cells (Tregs) which function in parallel with exhaustion to dampen potentially harmful inflammatory responses. Indeed, molecular analyses reveal that TFH and Treg gene expression patterns partially overlap with that of exhausted CD4 T cells.^66,71^ Our findings here only present an association between PD-1 expression, antigen-experienced T cell populations, and the chronic disease state of disseminated Coccidioidomycosis which presents an opportune milieu to induce exhaustion. More extensive molecular interrogation of Coccidioides-specific CD4 T cells in uncomplicated and disseminated disease is necessary to define exhaustion in this setting.

Most patients with DCM do not have a clearly identified immune defect to explain their failure to control disease. We showed here that many of these subjects have deficient T cell responses which may be augmented by adjunct immunomodulation. Immunotherapy for Coccidioides was first described in the 1970s with the use of Transfer Factor, which reportedly led to clinical improvement and augmentation of lymphocyte responses.^72,73^ IFNγ then emerged in the 21^st^ century for patients with or without a range of genetic defects.^74^ Dupilumab, which blocks IL-4 signaling to enhance Type 1 T cell responses, is effective for patients that exhibit a Th2-skewed phenotype.^14^ The success of these approaches emphasizes the importance of further investigation into immunologic deficits underlying anemic T cell responses in DCM to identify new therapeutic targets. PD-1 blockade has shown promising results in case reports of invasive mold infections, and may have broader applications in treatment refractory or severe infectious diseases.^33–37^ Our results suggests that immune checkpoint inhibitor therapy may be a valuable therapeutic approach for some individuals with DCM and deserves further investigation.

## Methods

### Ethics Approval

All patients provided informed consent to participate in protocols approved by the Institutional Review Board (IRB) of the University of California, Los Angeles, with reliance agreements from the Valley Fever Institute and the University of California, Davis.

### Patient Recruitment and Demographic

Subjects were confirmed to have coccidioidomycosis by an infectious disease specialist from the Valley Fever Institute, University of California, Davis, or University of California, Los Angeles. Disease category as described by Krogstad et al was assigned by these experts after review of diagnostic testing and clinical trajectory. Subjects were then enrolled in an IRB-approved protocol and whole blood was collected and shipped to UCLA for analysis. Demographic data was provided by subjects at time of recruitment. Self-identified race and ethnicity (SIRE) was recorded separately as ethnicity (Hispanic or non-Hispanic) and ethnicity (Black, White, Asian, Pacific islander or Hawaiian, or American Indian) per 1997 US census guidelines. Age, sex, and date of Coccidioides diagnosis were similarly self-reported by subjects at recruitment.

### Sex as a biological variable

Subjects were recruited independent of sex, and sex was analyzed as an independent variable.

### Arthroconidia (FKC) Generation, Harvest, and Fixation

Coccidioides immitis RS was obtained from BEI (RS NR-48942). Arthroconidia was harvested as previously described.^75^ Briefly, loopful samples from frozen stocks were inoculated onto 100 ml of 2× Glucose Yeast Extract (2× GYE; 2% glucose Sigma-Aldrich, #G8270-5KG; 1% yeast extract, Gibco #DF210929) 1.5% agar (Gibco #DF0145-17-0) in T225 tissue culture flasks with 100X penicillin/streptomycin (pen/strep; 10,000 units/ml of penicillin and 10,000 µg/ml streptomycin, Gibco #15140122) and grown for 4-10 weeks at room temperature. Arthroconidia were harvested as described and stored at 4°C. Arthroconidia viability was assessed by plating 100 µl of serially diluted samples on 2× GYE agar + pen/strep. Colony forming units (CFUs) were obtained by enumerating colonies after 3-4 days of incubation at 30°C. Fixed arthroconidia samples were generated by mixing arthroconidia aliquots with 37% formaldehyde in 1:5 ratio (6% final FA concentration) and incubating at room temperature for at least 30 minutes. Fixed arthroconidia were washed with 1× PBS twice, resuspended in 1× PBS (Gibco #10010049), and stored at 4°C.

### Spherule (FKS) Growth and Fixation

To generate C. immitis RS spherules, 1 × 106 arthroconidia/ml were inoculated into polypropylene vented shaker flasks (Fisher Scientific #BBV12-5) containing Converse medium^76^ and grown under shaking conditions (150 rpm) at 39°C supplemented with 10% CO2. After 5 days, mature spherules were observed. Spherules were fixed in a 1:5 ratio with 37% formaldehyde, incubated at room temperature for at least 30 minutes. The fixed spherules were washed with 1× PBS twice, resuspended in 1× PBS, and stored at 4°C. Spherules were quantified by direct counting using Kova hemocytometers (Fisher Scientific #22-270141).

### Cell Culture

Cells were cultured in complete T cell medium (RPMI 1640 supplemented with 10% FCS, 10 mM HEPES, 1 mM sodium pyruvate, 55 μM 2-mercaptoethanol, and 1X Pen-Strep) at 37°C.

For AIM assays, cryopreserved PBMCs were thawed at 37°C and washed twice with T cell medium. Cells were either cultured in media alone (No Stim) or stimulated with FKS (5,000 spherules per 1 million cells), FKC (1,000 conidia per 1 million cells), or anti-CD3-CD2-CD28 (StemCell ImmunoCult™, Cat #10970) for 24 or 48 hours before analysis by flow cytometry. MHC-I blockade was performed with 10ug/mL anti-HLA A,B,C clone W6/32 (Biolegend Cat #311427). MHC-II blockade was performed with 10ug/mL each of anti-HLA-DR, DP, DQ clone Tü39 (BD, Cat #555556), and anti-HLA-DR clone L243 (Biolegend, Cat #307665). Monoclonal antibodies for cell culture were low endotoxin, azide-free.

For intracellular cytokine staining, PBMCs were thawed and washed as above, and stimulated with FKS and FKC for 48 hours as above. Brefeldin A (Biolegend, Cat #420601) was added for the final 12-24 hours of culture. Where indicated, 10 ug/mL anti-PD-1 (BioXCell, Cat #SIM0003) or IgG4 Isotype control (BioXCell, Cat #CP147) were added.

### Flow Cytometry

For phenotyping, fresh or frozen PBMCs were washed with FACS buffer (PBS with 2% FCS, 1nM EDTA), stained with a cocktail of conjugated monoclonal fluorescent antibodies for 20 minutes at 4°C, and then washed twice with FACS buffer prior to data acquisition. Cultured cells from AIM assays were stained by the same protocol. ICS samples were fixed with 2% paraformaldehyde in PBS prior to surface staining, then permeabilized with Biolegend perm buffer (Biolegend, Cat #421002). Intracellular cytokine staining was performed in perm buffer at 4°C for 35 minutes. Cells were then washed twice with perm buffer and twice with FACS buffer before data acquisition.

Stain panels as follows. T cell Phenotype: CD3 BV605 (Biolegend, Cat #317322), CD4 PerCPCy5.5 (Biolegend, Cat #350008), CD8 PE (Biolegend, Cat #301051), CD45RA BV421 (Biolegend, Cat #304130), CCR7 PECy7 (Biolegend, Cat #353414), CD57 BV785 (Biolegend, Cat #393330), PD-1 AF647 (Biolegend, Cat #329910), HLADR AF488 (Biolegend, Cat #327010). AIM assay: CD3 BV605, CD4 PerCPCy5.5, CD8 PE, CD69 AF488 (Biolegend, Cat #310916), CD134 APC (Biolegend, Cat #309818), CD137 PECy7 (Biolegend, Cat #309818). ICS: CD3 BV605, CD4 PerCPCy5.5, CD8 PECy7 (Biolegend, Cat #344712), IFNγ PE (Biolegend, Cat #502509), IL-17 AF647 (Biolegend, Cat #512310), IL-4 BV421 (Biolegend, Cat #500826). Data were collected on a Cytek DxP10 digital flow cytometer and analyzed with FlowJo software.

### Statistical analysis and Data Handling

For flow cytometry results, AIM gates were drawn for each experiment using the unstimulated negative controls and anti-CD3-stimulated positive controls as guides. ‘AIM positive’ values for each assay were determined by subtracting unstimulated background from the FKC- or FKS-stimulated cells within each subject. Samples with less than 2,000 viable CD4 T cells upon data acquisition were discarded. We set a cutoff value of 0.05% for background-subtracted values to be determined as positive: values below this were considered negative. Over the course of this study, AIM assays were performed with FKS or FKC for 24 or 48 hour intervals due time, reagent, and sample constraints. We compiled results into a ‘composite’ AIM score which reports the average of positive results from assays performed for that subject. We determined that 24 or 48 hour stimulation with FKC or 48 hour stimulation with FKS were sufficient to detect Coccidioides-specific T cell responses and gave concordant results, but 24 hour FKS stimulation was not (**Figure 2**). Subjects with only a negative 24-hour FKS result were therefore not included.

Categorial variables were analyzed by Chi-square or Fisher exact tests. T cell phenotype results were assessed for normal distribution by visual inspection and analyzed by two-tailed nonparametric tests (Wilcoxon/ Mann-Whitney). Variation of phenotypic markers by age were assessed by simple linear regressions. AIM and ICS assay results were analyzed by paired or unpaired nonparametric Wilcoxon tests. Correlations were analyzed by nonparametric Spearman tests. Odds ratio confidence intervals were calculated by Baptista-Pike method. Corrections for multiple tests were not performed. Statistical analyses were performed in R and Graphpad Prism.

## Data Availability

Values for all data points in graphs are reported in the Supporting Data Values file.

## Author Contributions

Conceptualization: GDW, TJT, MIG-L, MJB

Investigation: GDW, TJT, AVS, MAML

Fungal antigen preparation: MMT, SB

Resources (patient and samples): GRT, RHJ

Visualization: GDW

Funding acquisition: MJB

Writing (original): GDW

Writing (editing): all

