## Supplemental for "Exhausted T cell phenotypes in disseminated coccidioidomycosis"

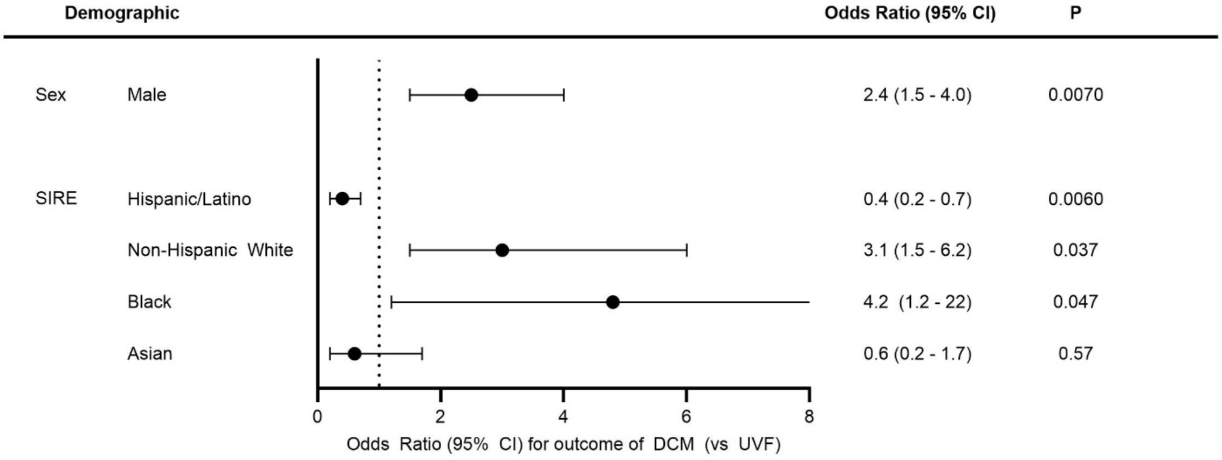


**Figure S1: Forest plot showing association of sex and SIRE (self-identified race and ethnicity) with disease severity**

Odds ratios of Male sex (relative to female sex) and SIRE of Hispanic (relative to non-hispanic), Non-Hispanic White (relative to non-white), black (relative to non-block), and asian (relative to non-asian) for outcome of DCM vs UVF. Odds ratios by Baptista-pike, p-values by Fisher Exact test.

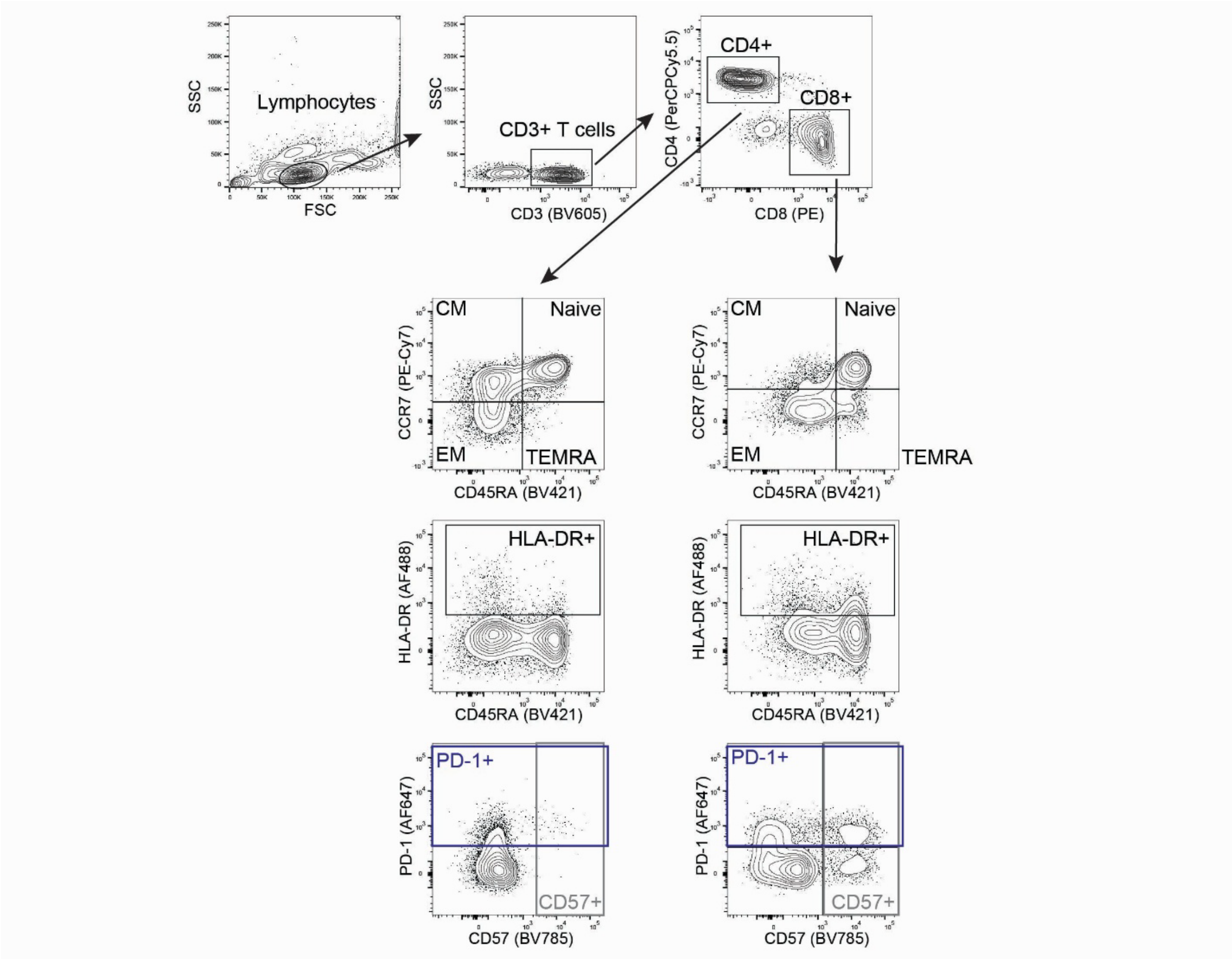


**Figure S2: Representative PBMC T cell Phenotype Gating Strategy** (related to **Figure 1**)

Flow cytometry staining of cryopreserved PBMCs was performed as described in methods. CM, central memory; EM, effector memory.

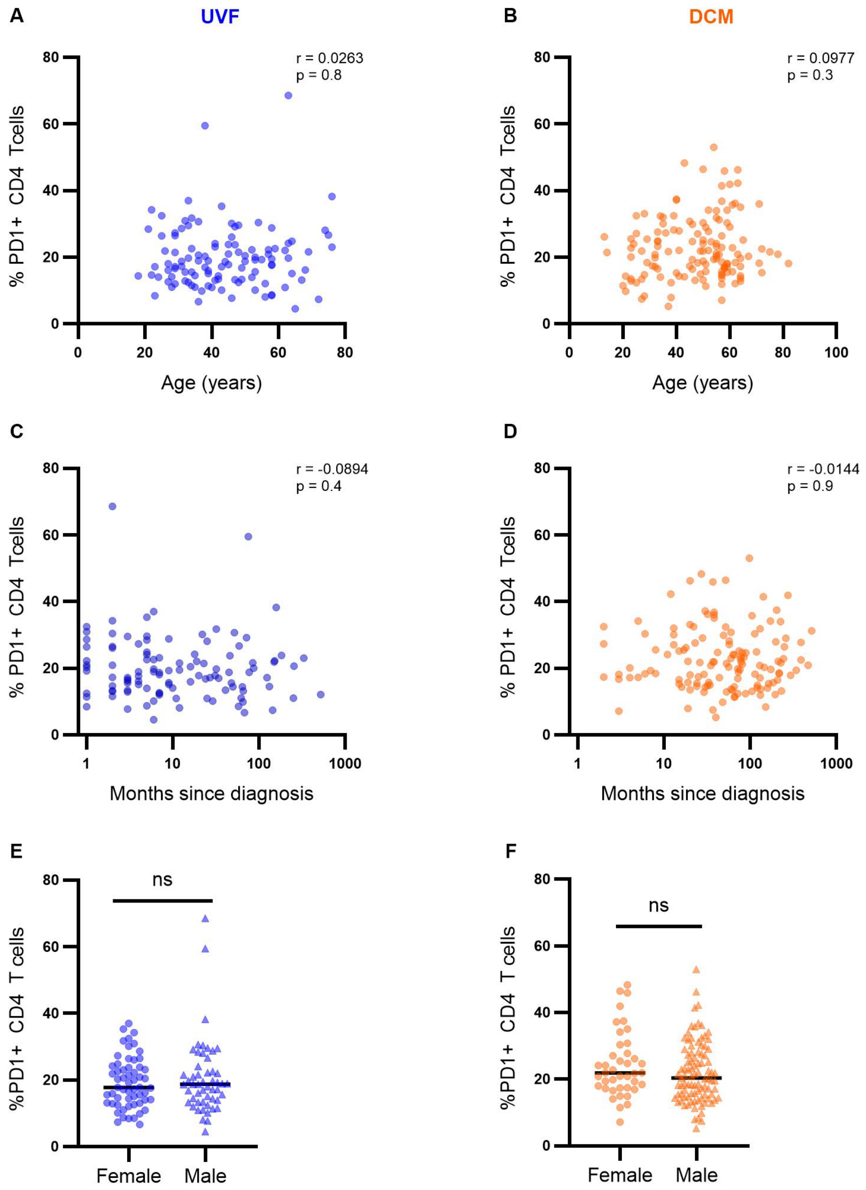


**Figure S3: Association of CD4 T cell PD-1 expression with age, time since diagnosis, and sex in UVF (**blue, N = 109**) and DCM (**orange**,** N = 135**)**, (related to **Figure 1**)

(**A**, **B**) Spearman correlation of %PD1+ CD4 T cells with age. (**C**, **D**) Spearman correlation of %PD1+ CD4 T cells with time since diagnosis in months. (**E**, **F**) Mann-Whitney test comparing %PD1+ CD4 T cells by sex (Female and Male). ns, not significant

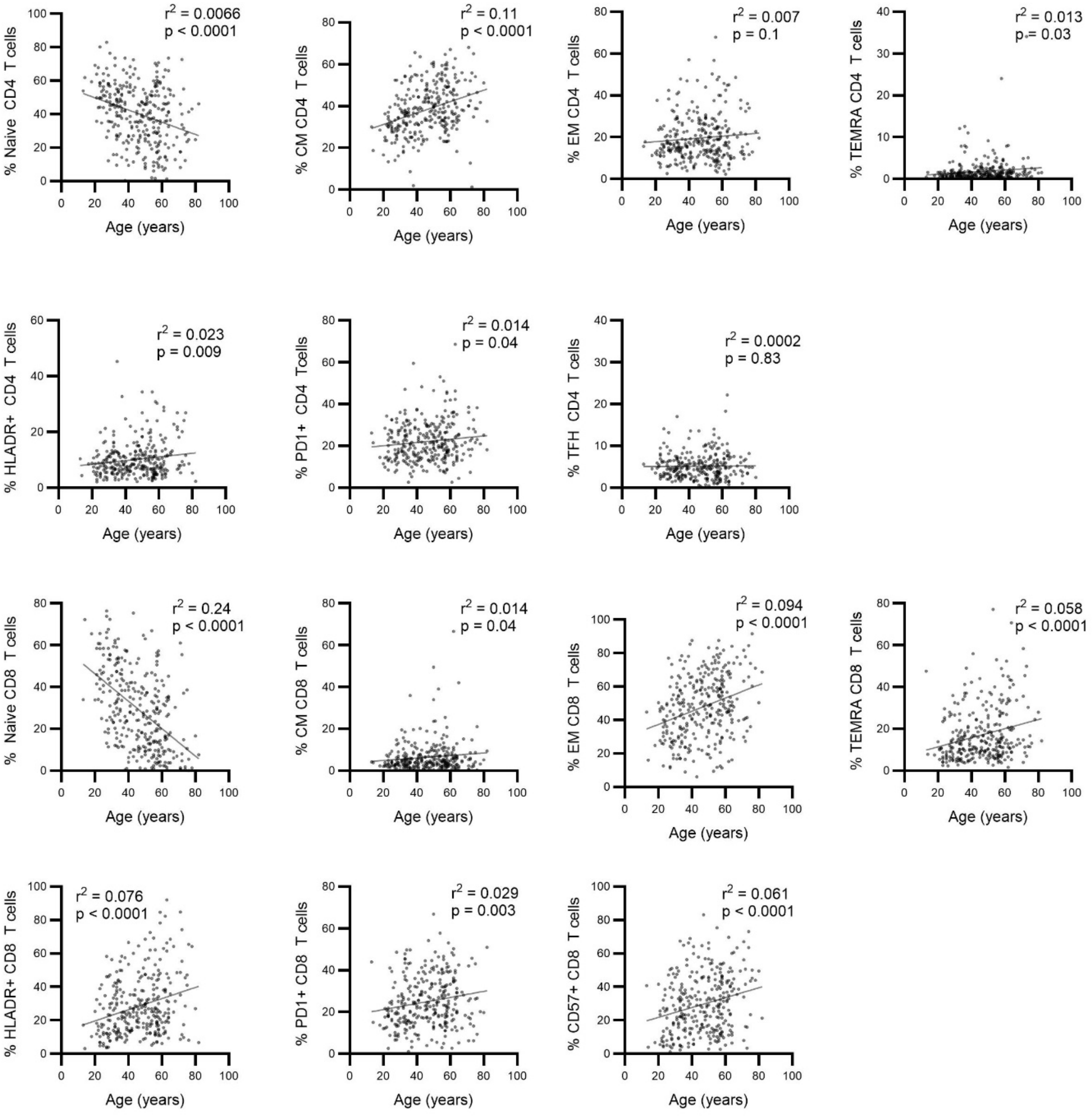


**Figure S4: Variance of phenotypic markers with age** (related to **Figure 1**)

Simple linear regression was performed for each phenotyped population on CD4 and CD8 T cells by subject age, $r^2$ and unadjusted p values are presented. N = 302 subjects.

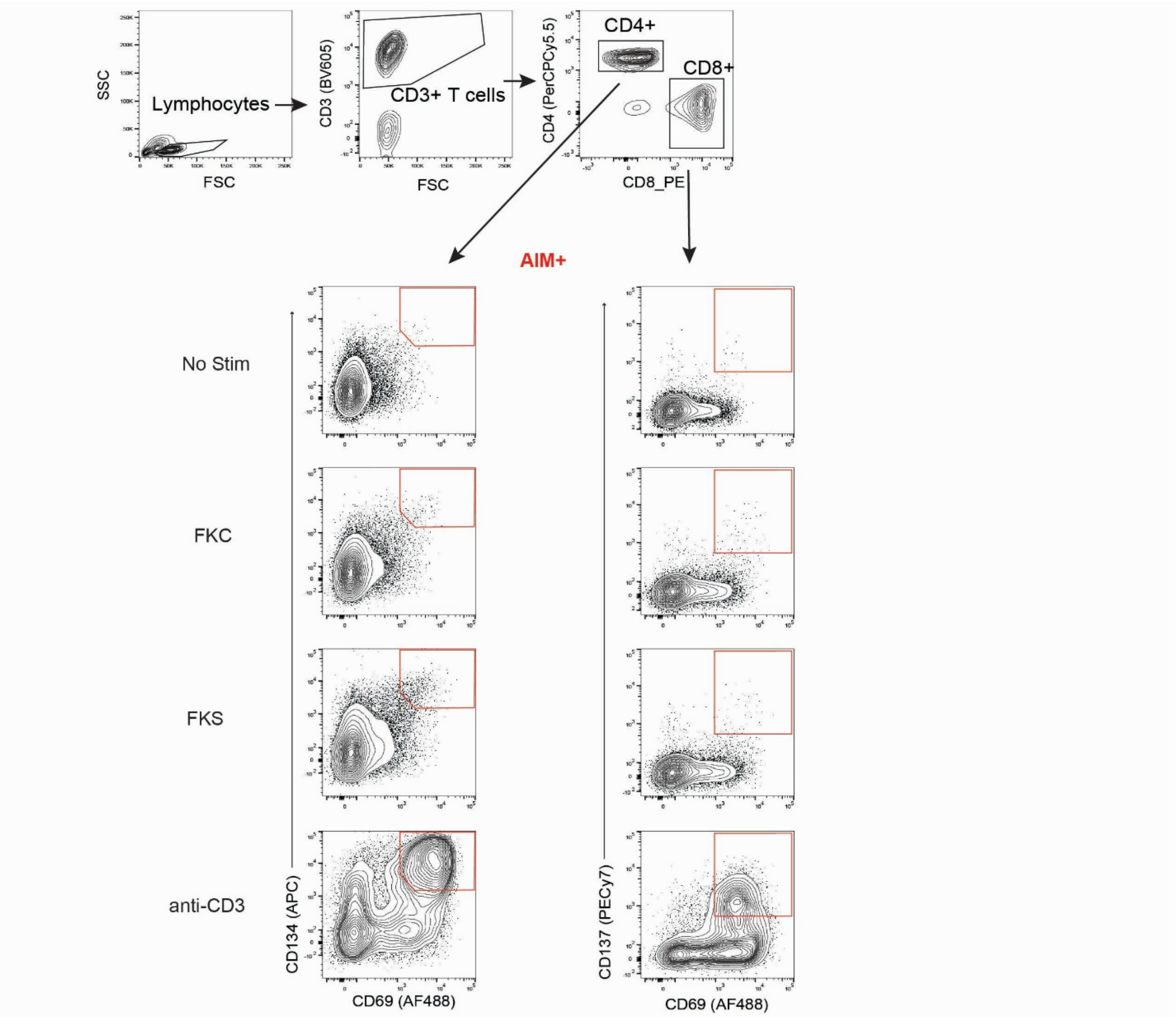


**Figure S5: Representative AIM Gating Strategy (**related to **Figure 2)**

Flow cytometry staining of cultured PBMCs performed as described in methods. AIM positive population (red) was determined as the CD69+CD134+ subset of CD4 T cells and CD69+CD137+ subset of CD8 T cells following culture of PBMCs alone (No Stim) or stimulation with FKC, FKS, or anti-CD3. The AIM positive population was gated conservatively to minimize background

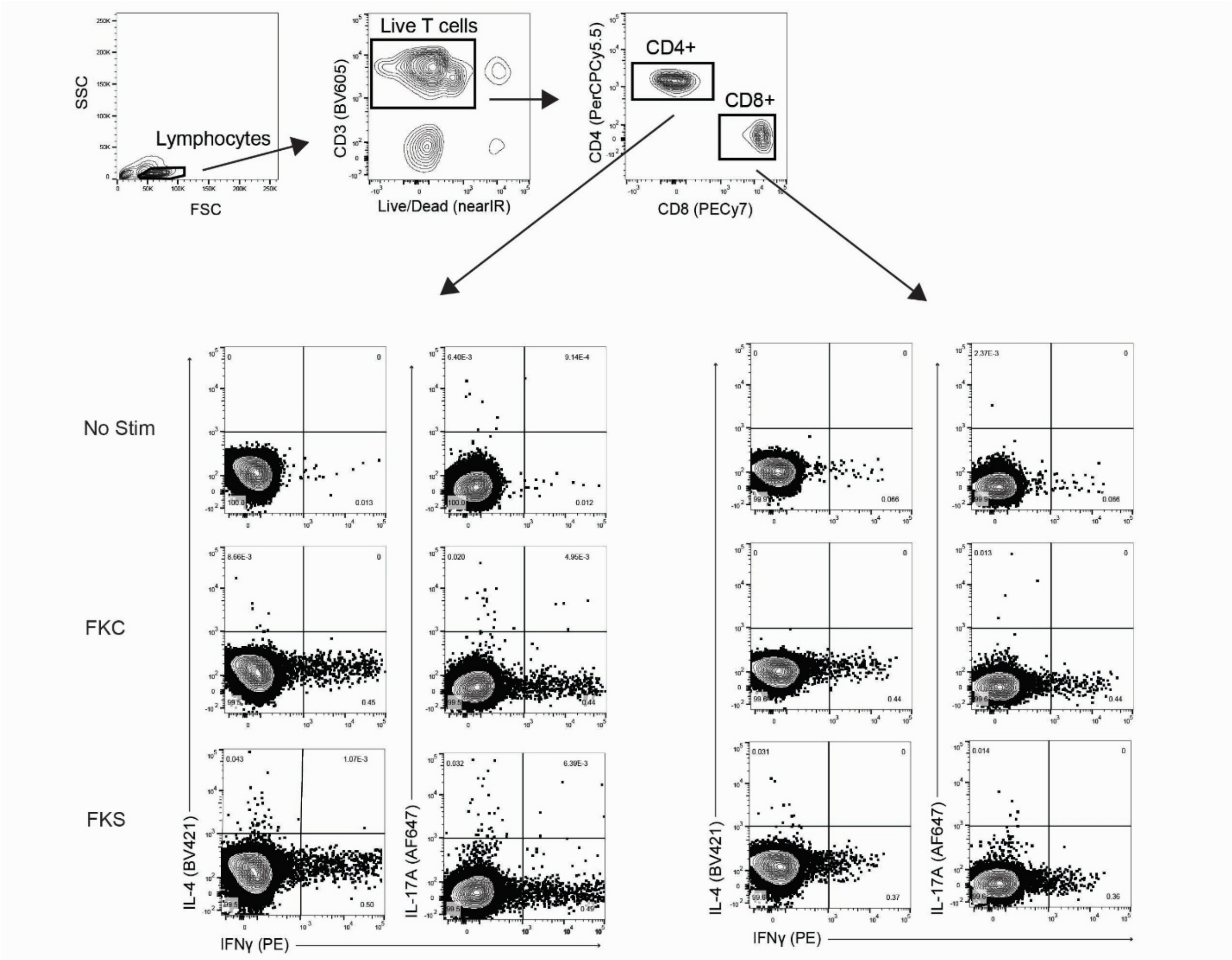


**Figure S6: Representative ICS Gating Strategy (**related to **Figure 3, 4)**

Intracellular staining of cultured cells was performed as described in methods. Representative gating of IFNγ, IL4, and IL17A positive cells by quadrants for CD4 and CD8 T cells is shown. Only CD4 T cell results were included in analysis as CD8 T cell responses were MHC-I-independent and may represent bystander activation (as presented in Figure 2).

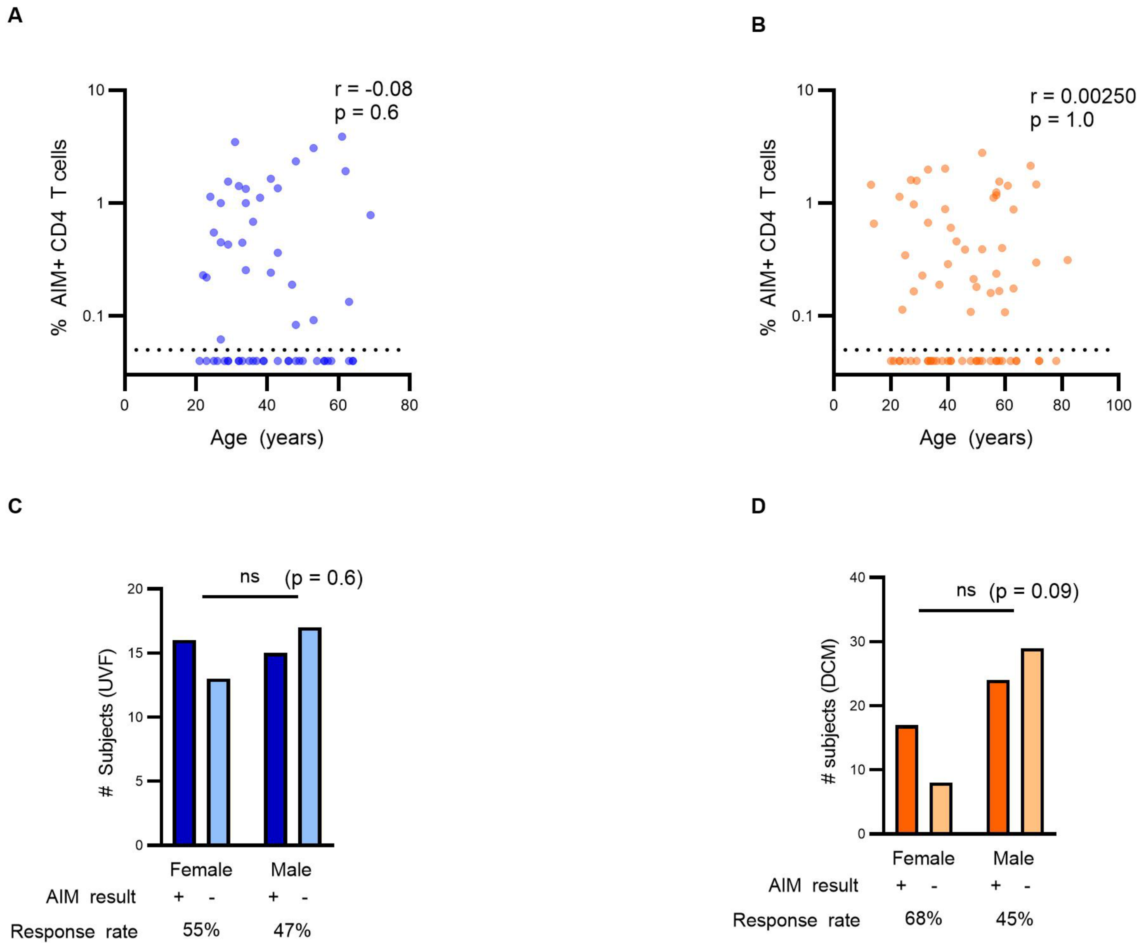


**Figure S7: Association of AIM results with Age and Sex (**related to **Figure 3)**

(**A**, **B**) Spearman correlation of age with %AIM+ CD4 T cells for (**A**) UVF (N = 31) and (**B**) DCM (N = 40). (**C**, **D**) Fisher exact tests comparing AIM results (positive/+ or negative/-) among male and female subjects within (**C**) UVD and (**D**) DCM. ns, not significant, p > 0.05.

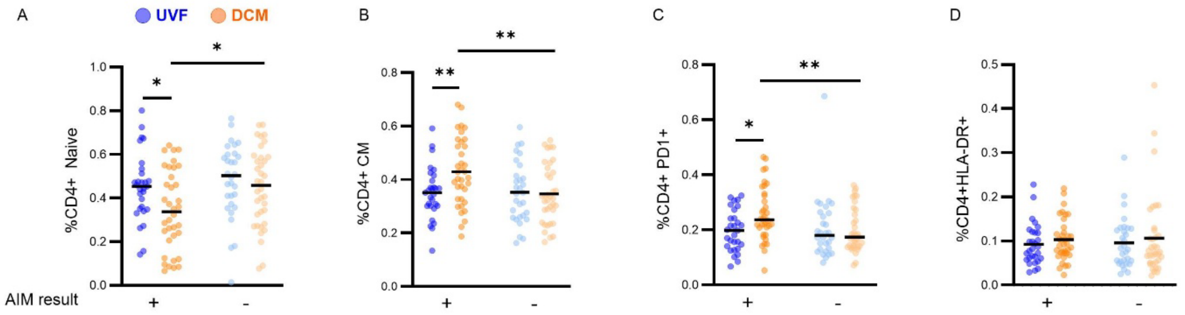


**Figure S8: Differences in T cell phenotype between AIM positive (+) and AIM negative (-) UVF (blue) and DCM (orange) subjects (**related to **Figure 4)**

PBMC CD4 T cell phenotypes (as presented in Figure 1) were compared between AIM-positive UVF (dark blue, N = 29), AIM-positive DCM (dark orange, N = 34), AIM-negative UVF (light blue N = 30), and AIM-negative DCM (light orange N = 39) subjects. We specifically assessed (**A**) CD4 Naïve T cells, (**B**) CD4 central memory (CM) T cells, (**C**) CD4+ PD-1+ T cells, and (**D**) CD4 HLA-DR+ T cells. Statistical comparisons by Mann-Whitney U test. *, $p < 0.05$; **, $p < 0.01$.

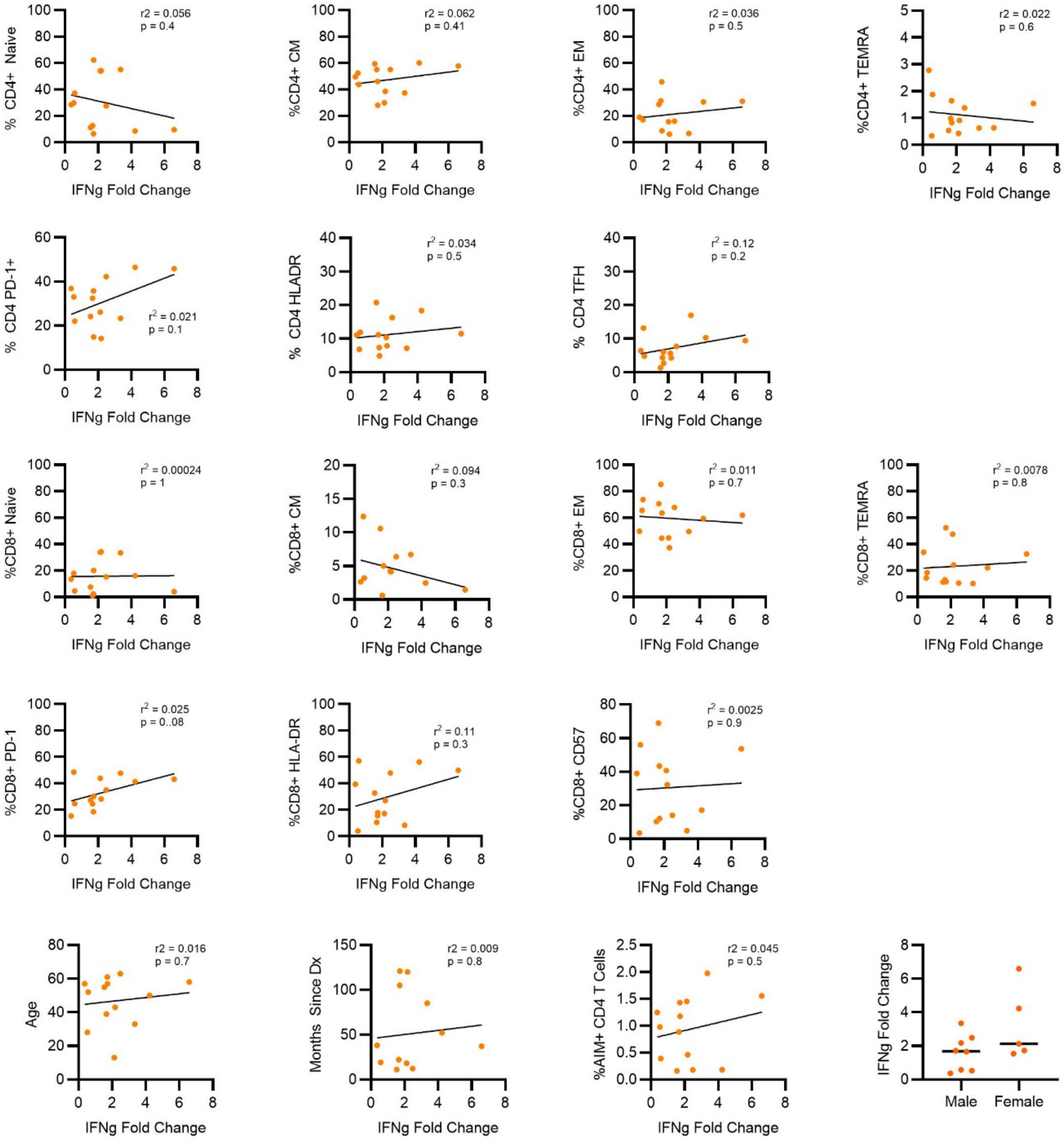


**Figure S9: Variance of response to checkpoint blockade with T cell phenotypes and demographic variables**

Simple linear regression was performed for fold change interferon gamma production in response to Coccidioides antigen stimulation (IFNg fold change) compared to T cell phenotypic markers and demographics. $r^2$ and unadjusted p values are presented. Variation by sex compared by Mann Whitney U test. No significant variance was found. N = 13 subjects.
